# Identification of Global DNA Methylation Signatures in Patients of High Altitude Induced Venous Thrombo-Embolism (HA-VTE)

**DOI:** 10.1101/2022.03.27.22272933

**Authors:** Swati Srivastava, Iti Garg, Babita Kumari, Uday Yanamandra, Jasjit Singh, Lilly Ganju, Rajeev Varshney

## Abstract

**Background:** Pathophysiology of venous thrombo-embolism (VTE) depends upon several acquired, inherited and environmental risk factors, including high altitude (HA) exposure. The present study aims to gain insights into pathophysiological mechanism(s) of high altitude induced VTE (HA-VTE) by studying global methylation signatures.

**Methodology:** Blood samples were collected from Indian Army volunteers divided into four study groups; sea level control (SLC), sea level VTE patients (SL-VTE), high altitude control (HAC) and high altitude VTE patients (HA-VTE). Methylation patterns were studied using whole genome bisulfate sequencing. Differentially methylated genes and pathways were identified by comparing percentage methylation.

**Results:** Highest DM was observed in SL-VTE (1162 gene) compared to SLC where in hyper methylation was predominant (62.9%) compared to hypo methylation (37.05%). A reverse trend was observed in HA-VTE, where hypo methylation (61.69%) was predominant over hyper methylation (38.30%) in a total of 296 DM genes. Differential hypomethylation of genes involved in cell adhesion/platelet activity (CADM1, PTPRK, PDGFA) and immune response (CXCL12, IL4, IRF4, NLRP1) was observed in HA-VTE whereas genes encoding transcription factors (GSC, RPSKA1), trans membrane receptor (NOTCH2) and growth factor (TGFB2) were hypermethylated in comparison to SL-VTE. Methylation pattern of HA-VTE compared to HAC showed hypomethylation in genes involved in oxidative phosphorylation (CPOX), immune response and stress response (NDRG1), while those involved in signaling mechanisms (KALRN), neurotransmitter release (TMPRSS2) and transcription factor (ELF1) were hyper-methylated.

**Conclusions:** Our study for the first time reveals genome wide methylation pattern in HA-VTE group where in differential hypo methylation in cell adhesion and inflammation genes was observed.

## Introduction

Thrombo-Embolic Disorders (TED) comprising of both venous and arterial thrombosis together results in significant morbidity and mortality every year worldwide. Thrombosis is the third most common underlying pathology of major cardiovascular complications like ischemic heart disease and stroke, and remains a major contributor to the global disease burden (Raskob et al., 2014).The etiology of venous thrombo-embolism (VTE) is considered multi factorial, as the genetic and acquired risk factors individually or together commence the thrombus formation.VTE encompasses two clinically interrelated conditions; deep vein thrombosis (DVT) and pulmonary embolism (PE), the later one being potentially fatal. DVT most commonly starts in the leg, although it rarely also occurs in other veins such a supper extremities, liver, cerebral sinus, retina and mesenteric. The thrombus or blood clots might dislodge from the site of origin and travel through blood to lodge itself into lungs, leading to PE. One in every three patient of VTE manifests PE, together with DVT; while two in every three patient manifest only DVT (White, 2003). Several independent studies have established the role of genetic risk factors in VTE pathophysiology. These include, gene involved in fibrinolytic pathway such as tissue-type plasminogen activator (t-PA) and plasminogen activator inhibitor-1 (PAI-1) (Asselbergs et al.2007); deficiencies of natural anticoagulants as well as elevated levels of certain coagulation factors such as factor VIII and factor XI (Ageno et al. 2008, Cushman et al. 2009, Ryland et al.2012, Ota et al. 2011). Limited studies have been conducted on gene expression changes in VTE under hypoxic conditions (Jha et al. 2018, Srivastava et al. 2020).

‘Hypoxia ‘as encountered at high altitude (HA environment is associated to low oxygen bio availability and decreased ambient pressure along with other environmental stressors like cold, radiation and compromised physiological conditions. Increasing evidence has demonstrated that environmental conditions prevailing at HA such as hypoxia, dehydration, heme concentration, use of constrictive clothing and enforced stasis because of severe weather may result in VTE event (Cancienne et al. 2017). Un-acclimatized rapid ascent to HA modulates coagulation parameters and hemostatic profiles (Singh et al., 1972, Doughty et al., 1994) and incidences of DVT and PE are reportedly higher in sojourns travelling to HA and soldiers posted at HA without any other co-existent risk factor (Kumar 2006, Anand et al. 2001, Khalil et al.2010, Rathi et al. 2012).

Epigenetic modifications such as DNA methylation, histone modification and small non-coding RNA regulate gene expression by altering chromatin structure and accessibility, as well as interacting with DNA binding proteins on the regulatory regions. Gene expression is thus affected on account of the hindrance caused on the promoters/enhancers for the binding of transcriptional factors, activators/repressors (Chamberlain et al., 2014). DNA methylation marks the interface between genetic and environmental risk factors of complex human diseases. Cells are programmed to respond towards environmental stimuli, one of which is hypoxia. Various evidences suggest that regulation of gene expression by hypoxia has epigenetic basis, especially DNA methylation. It is a dynamic epigenetic modification and crucial for regulating gene expression, genomic imprinting and is the defensive mechanism against chromatin instability (Putiri et al. 2011, Suzuki et al. 2008). Tumor hypoxia is suggested to be a novel regulator of DNA methylation as tumour suppressor gene promoters in tissue from cancer patients were found to be highly methylated in hypoxia (Thienpont et al., 2016). Also, the data strongly suggest that up to half of hyper methylation events were attributable to hypoxia (Thienpont et al., 2016). Hypoxia induced decrease in gene transcription has epigenetic basis as reviewed by Perez-Perri and coworkers. They further provided multiple compelling evidences to demonstrate chromatin changes such as DNA methylation and histone modifications during HIF activation under hypoxic conditions (Perez-Perri et al. 2011).

Systematic assessment and analysis of global methylation pattern an help in better understanding of effect of changing environment in VTE pathophysiology. Environmental and stress related challenge ssuch as life style habits, air pollutants etc. alter another wise un-methylated chromosomal region and changes the DNA methylation pattern (Baccarelli et al.,2009; Bollati et al., 2010; McGowan et al., 2009). Substantial evidence demonstrates that DNA methylation is associated to hypoxia as well as cardiovascular diseases. Aberrant promoter hyper methylation is documented in atherosclerosis, coronary artery disease etc. (Friso et al.2012, Yamada etal., 2018), however, the role of global DNA methylation in HA induced coagulation as well as in DVT is yet to be explored. DNA methylation marks were found to be strongly associated with coagulation factor V Leiden (FVL) mutation (Aissi et al.,2014).Moreover, DNA methylation patterns have been found by global DNA methylation analysis of anti phosphor lipid syndrome patients, suggesting its stronger link to thrombotic disorders (Kim etal.2012).

In summary, whether hypoxia has an underlying methylation mechanism to modulate thrombotic propensity is yet to be elucidated. No direct evidence is available on alteration of DNA methylation machinery during VTE event under influence of hypoxia. Present study has been conducted on Human subjects in order to understand the role of aberrant DNA methylation, regulating thrombotic events during high altitude exposure. Our objective was to gain insights into global DNA methylation status of patients of high altitude induced venous thrombo-embolism in relation to patients at sea level.

## Methodology

### Study subjects

The present study involved Human volunteers and was conducted in accordance with the ethical guidelines of Indian council of Medical Research after approval from institutional human ethical committee. All volunteers recruited for the study signed an informed consent for their participation. Venous thrombosis patients were from Indian Army Soldiers and their samples were collected from the Army Research and Referral Hospital (R & R), Delhi, India (sea level VTE patients) and the Western Command Hospital, Chandi Mandir, Chandigarh (high altitude VTE patients). Patients with history of pre-existing systemic disease, any prior surgery, vasculitis and malignancy were excluded from our study. Also, clinical profile of patients, time and site of venous thrombosis (VT) episode along with the presence of predisposing factors (such as surgery, trauma, prolonged immobilization, hypertension, diabetes, familial history of bleeding) were documented at the time of sample collection. Diagnosis of VTE was confirmed for all patients by atleast one of the radiological imaging methods such as colour Doppler/contrast-enhanced computed tomography/computed tomography angiography/magnetic resonance imaging. Samples from age and sex matched healthy Army soldiers posted at Delhi and those deployed at high altitude region (Leh, Ladakh) were taken as controls. The persons with history of any VTE incident were excluded from the control group.

### Case-Control Study: Subject groups and sample collection

A total of 18 study subjects were divided into four groups; high altitude VTE patients (HA-VTEn=5), high altitude controls (HAC n=5), sea level VTE patients (SL-VTE n=4) and sea level controls (SLC n=4).

Sodium bisulfate sequencing was used for quantitative DNA methylation measurements. Approximately 1 ml of blood from each volunteer was collected in EDTA tube. Demographic data recording for each subject was also do neat the time of sample collection.

### DNA isolation and quantitation

Genomic DNA was isolated from peripheral blood using QIAamp DNA isolation kit (Qiagen, Germany), according to manufacturer’s instructions. Quantitative analysis of high molecular weight DNA was done using DNA/RNA nano drop 2000 spectrophotometer (Thermo fischer, USA). DNA was also assessed qualitatively on 0.7% agarose gel containing ethidium bromide.100 ng of DNA was loaded in each well and run on 50V for 30 min and visualized under UV.

### Library preparation and Bisulfite sequencing

For library preparation, DNA samples were fragmented and specialized adapters were added to each end. These adapters contain complementary sequences that allow the DNA fragments to bind to the flow cell (Illumina methylation kit). Bisulfite conversion changed unmethylated cytosines to uracil during library preparation. In bisulfite sequencing (BS-Seq), denatured DNA was subjected to bisulfite treatment, which converted cytosine (C) residues to uracil (U), but left5-methyl cytosine residues unaffected. Converted bases were identified (after PCR) as thymine

(T) in the sequencing data. These DNA fragments were then amplified and purified. The DNA library was loaded onto the flow cell and placed on a sequencer (service obtained from M/s Sandor Life sciences Pvt. Ltd.).

### Sequencing and alignment

Bisulfite conversion of genomic DNA followed by next generation sequencing (BS-seq) is most widely accepted technique for measuring methylation state of whole genome at single base resolution. Paired end sequencing (150nt) was done using Illumina HiSeq platform. All 150 base pair, paired end reads were quality checked for low quality bases and adapter sequences. After processing of raw reads, quality check was done using Perl scripts. Because of C to T conversion, bisulfite sequence reads are not complementary to the reference genome and thus the processing of sequencing data is very challenging. Thus special alignment tool, Bismark was used for alignment of reads to reference genome. This tool completed the process in three steps;

(i) Genome preparation, (ii) Alignment and (iii) Methylation extractor. Bismark performed alignments of bisulfate treated reads to reference genome and cytosine methylation calling simultaneously. Mapping of bisulfate treated reads was done using short read aligner Bowtie 2. Sequence reads were first transformed into fully bisulfite-converted forward (C->T) and reverse (G->A) reads, before they were aligned to similarly converted versions of the genome (also C->T and G->A converted). Sequence reads that produced a unique best alignment from the four alignment processes against the bisulfite genomes were then compared to the normal genomic sequence and the methylation state of all cytosine positions in the read are inferred. A read was considered to be aligned uniquely if an alignment had a unique best alignment score. If a read produced several alignments with the same number of mismatches or with the same alignment score, then it was discarded altogether.

### Methylation analysis

Data obtained from methylation calling was used to calculate differential methylation (DM). Fisher’s exact test was used to calculate p-values. Differentially methylated regions/bases were then subjected to gene annotation using genomation package. In this package, the gene annotation from a BED file was read and annotated differentially methylated regions with that information using genomation functions. This detailed the exact percentage of differentially methylated regions on promoters/introns/exons/intergenic region. This was followed by comparative group analysis of hyper-methylated and hypo-mehylated regions amongst HA-VTE and SL-VTE patients with respective controls followed by pathway analysis and gene enrichment analysis.

The complete study work flow has been depicted in figure1.

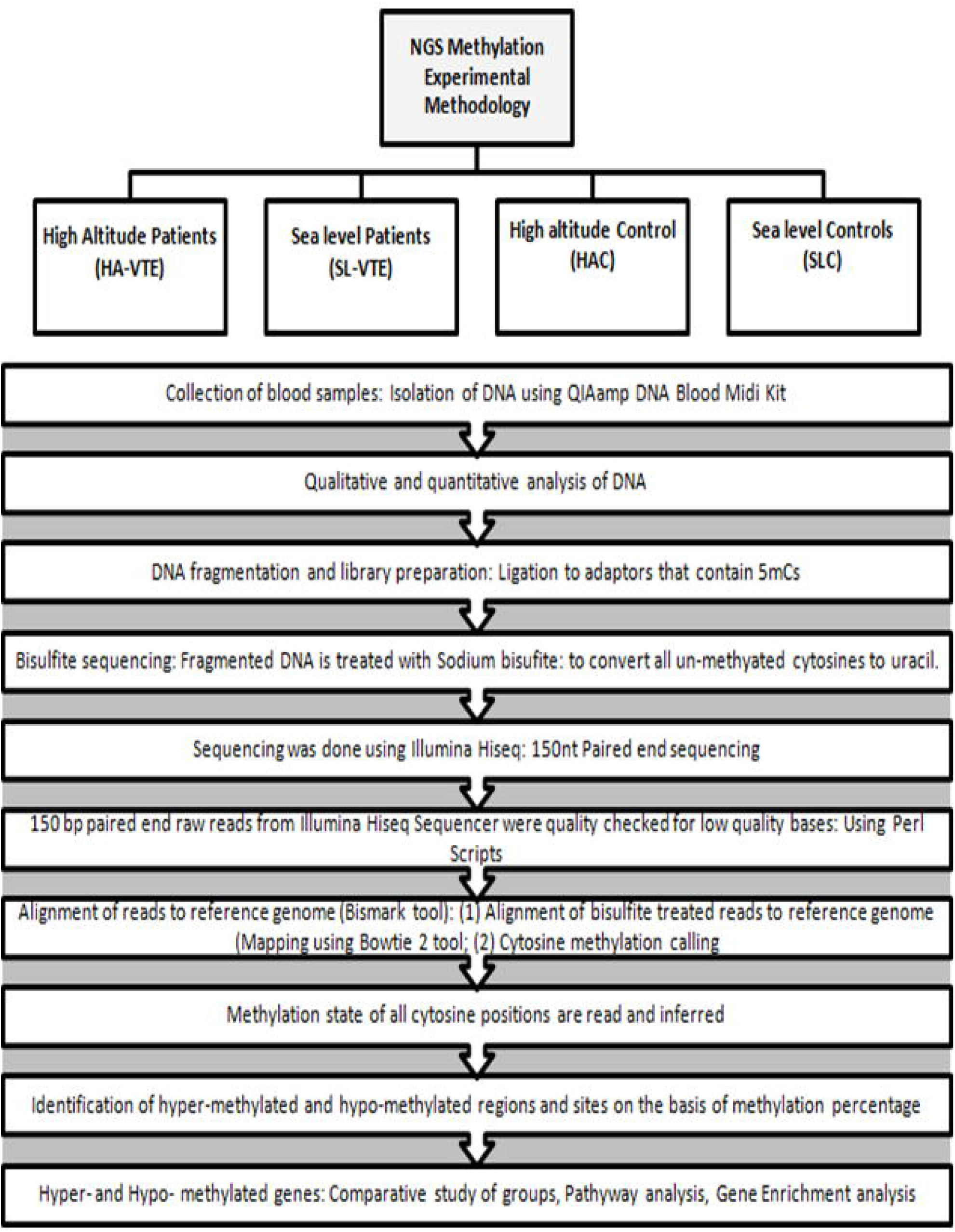

## Results

### DNA methylation features and Genomic features of DMR and DMS

Mapping efficiency of paired end reads was in between 78-80%. We studied DNA methylation widely in CpG (5’—Cytosine—phosphate—Guanine—3’) context as more than 80% of methylation events occurred at CpG sites. Methylation in CHG and CHH (where H correspond to A, T or C) context was only in between 0.1-0.2%. DM was studied in both differentially methylated regions (DMR) and differentially methylated sites (DMS). Percentage of methylation across different samples were observed based on those occurring in CpG islands, CpG shores, CpG shelves, transcription factor binding sites (TFBS) and in DNaseI hypersensitive sites (as detailed in figure 2a and 2b).Largest fraction of DMR and DMS was observed in DNase I hypersensitive sites (∼0.5 in DMR and 0.13-0.2 in DMS). TFBS has the next highest methylation proportion (0.3-0.4 in DMR and 0.07-0.09 in DMS). We observed a very small fraction of methylationin CpG shelves and CpG shores and least in CpG islands.

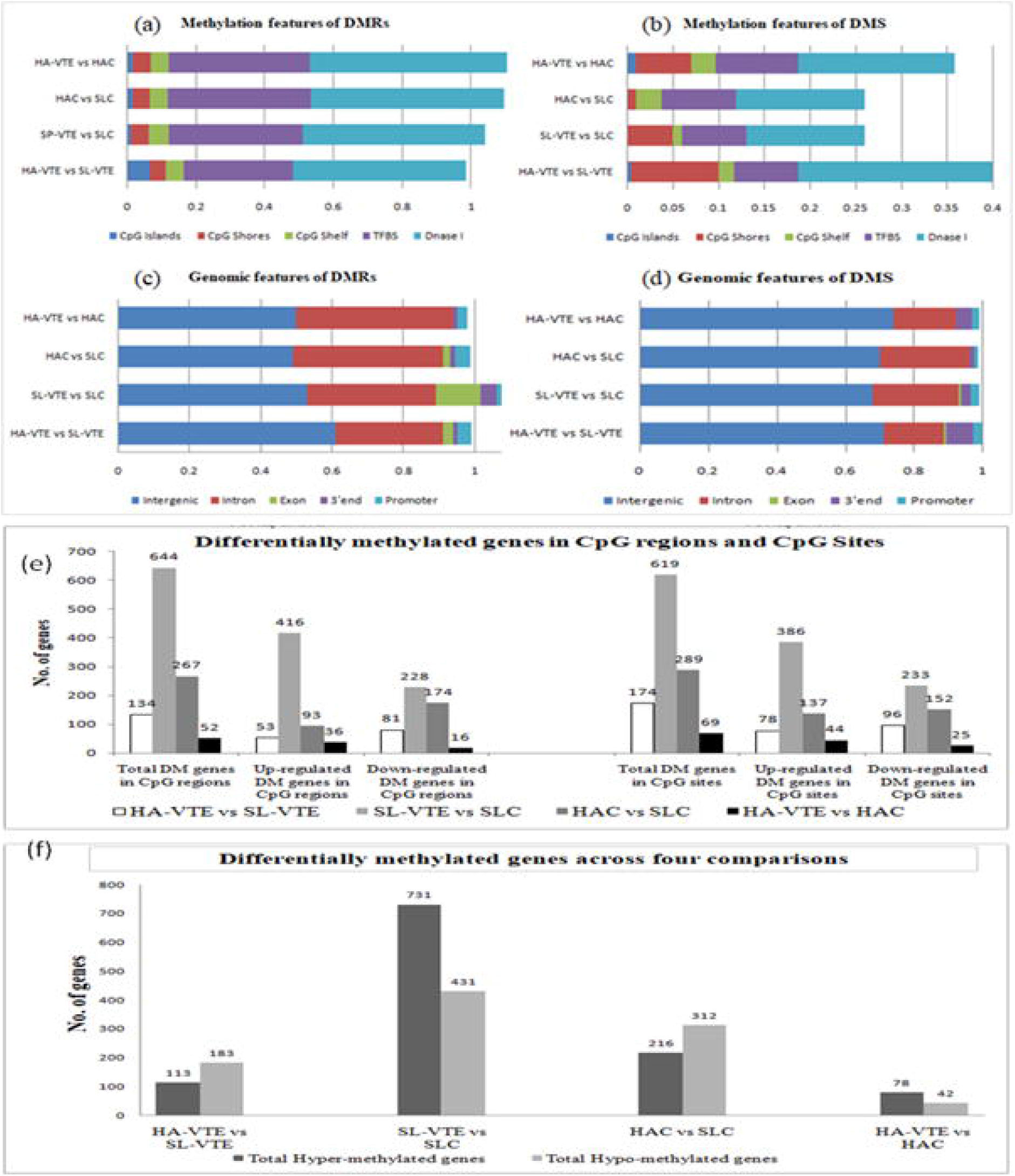

Regarding the genomic distribution of DMR and DMS, the percentage of methylation was studied in promoter regions, 3’end, exonic region and intronic region of gene and those occurring in intergenic region. We observed that 80-90% of methylation changes occurred in intergenic and intronic regions of the genes. A very small proportion of total differentially methylated regions/sites belonged to Exonic, 3’-region and promoter region of the genes (figure 2c and 2d).

### Identification of differentially methylated genes

Four types of comparative analysis was done amongst the different study groups; (i) HA-VTE vs SL-VTE; (ii) SL-VTE vs SLC; (iii) HAC vs SLC; (iv) HA-VTE vs HAC. Criteria for DM were≥ +25% for hyper methylation score and ≤ 25% for hypo methylation score. Highest number of CpG regions and CpG sites were observed in SL-VTE in comparison to SLC (figure 2e). SL-VTE vs SLC had highest number of hyper methylated and hypo methylated genes, wherein the component of hypermethylation was more (n=731) compared hypomethylation (n=431). In contrast, VTE patients at high altitude showed higher component of hypo methylation (n=183 genes) compared to hyper methylation (n=113 genes) in comparison to SL-VTE group. In case of HA exposed healthy controls (HAC) in comparison to SLC, number of hypo methylated genes were more(n=312) than hyper methylated genes (n=216). Least number of genes showed DM when HA-VTE were compared to HAC, only 78 genes showed hyper methylation whereas, 42 genes were hypo methylated (figure2f).A very small percentage of the methylation changes were observed in promoter region of genes. The distribution of DM in different genomics regions is illustrated in figure 3(a) and 3(b) using https://scatterplot.online/.

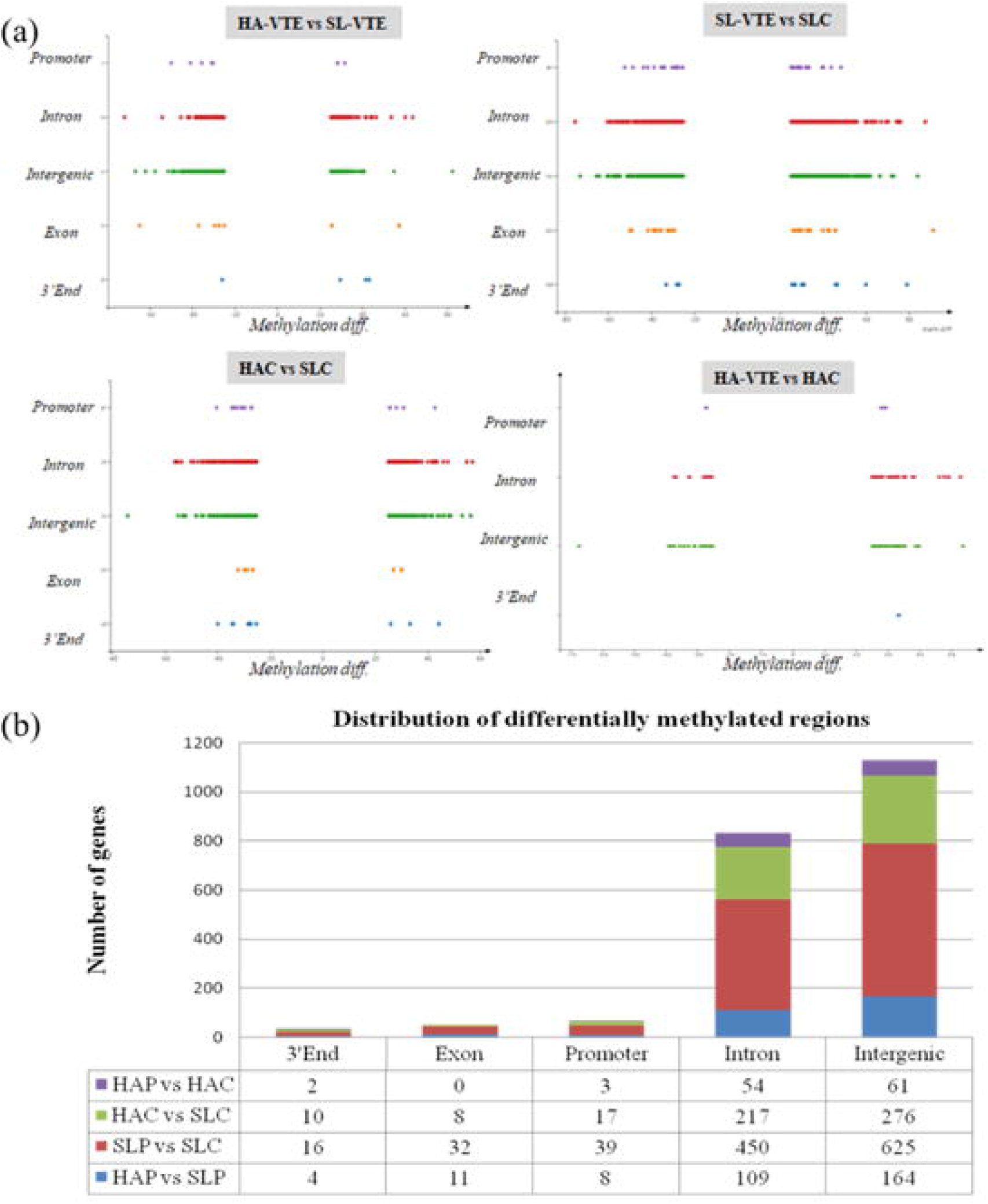

### Comparison of hypo-methylated and hyper-methylated genes across different study groups and Pathway analysis

Hypo methylated and hyper methylated genes from each comparison were compared with the help of Venn diagram (Figure4a and 3b) (Heberleetal.2015,http://www.interactivenn.net/). Maximum number of common differentially methylated genes was seen in SL-VTE vs SLC and HAC vs SLC (82 common hypo methylated genes and 80 common hyper methylated genes). These genes are detailed in figure 4c and 4d. Most of the genes common in other comparisons were either un-characterized or non protein coding. Protein coding differentially methylated genes for high altitude patients (as obtained from two comparisons; HA-VTE vs SL-VTE and HA-VTE vs HAC) and sea level patients (as obtained from comparison (SL-VTE vs SLC) are depicted in Figure 4.

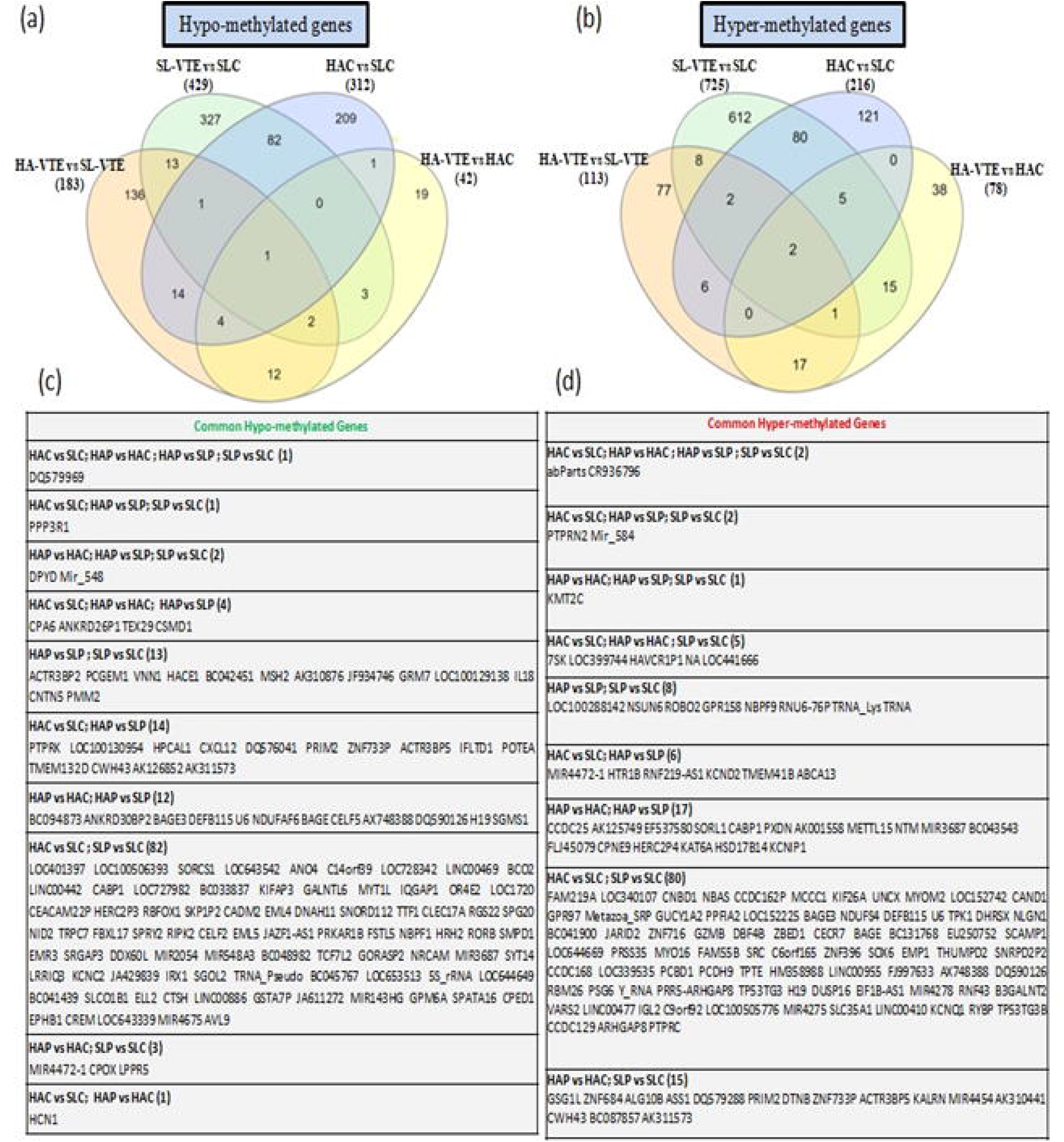

Pathway analysis was done using PANTHER pathways (http://pantherdb.org/tools/index.jsp) (Mietal.,2010) and REACTOME (https://reactome.org/PathwayBrowser/)(Croftetal.2014,Fabregatetal.2018) database. Differentially methylated (DM) pathways in HA-VTE in comparison to SL-VTE and HAC included AP-1 transcription factor network, β1 integrin cell surface interactions, endothelin signaling pathway, PDGF receptor signaling pathway, VEGF signaling, Thrombin/ protease activated receptor, urokinase-type plasminogen activator (uPA) and uPAR mediated signaling etc., which included both hypo methylated and hyper methylated genes (supplementary table 1and2). Apart from these, some hypo-methylated genes in HA-VTE attributed to EPO signaling pathway, HIF-1α transcription factor network, inflammasomes, heme biosynthesis, lipid metabolism etc. Amongst unique hyper methylated pathways were potassium channels, toll receptor cascades, Angiopoietin receptor or Tie1 mediated signaling, e-cadherin signaling events and GABA synthesis etc (Figure 5).

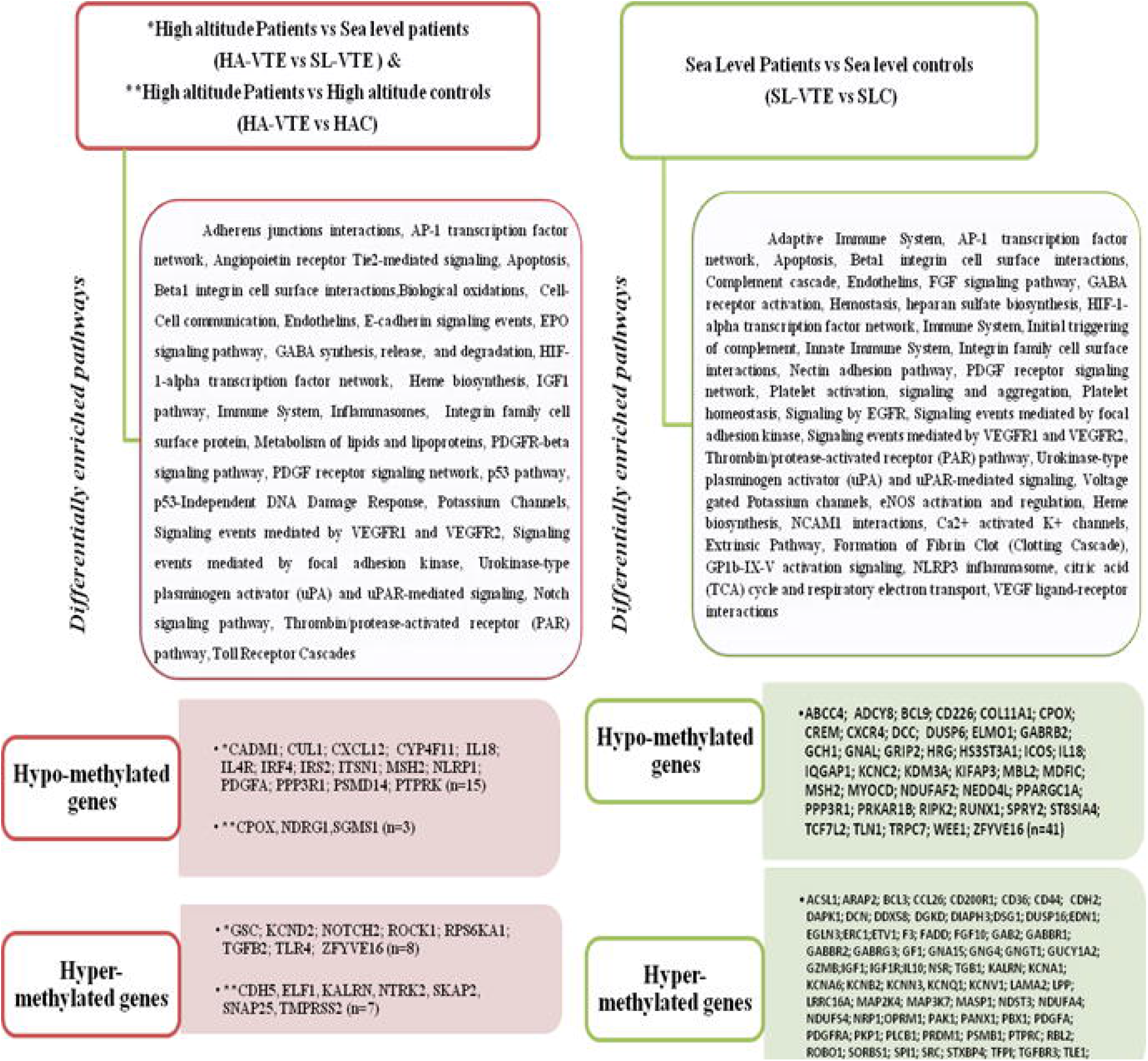

Number of DM pathways in SL-VTE was much more compared to HA-VTE. These included pathways with both hyper- and hypo-methylated genes viz., adaptive immune system, AP1 transcription factor network, apoptosis, β-1integrin cell surface interactions, complement cascade, endothelin signaling, FGF signaling, GABA receptor activation, hemostasis, HIF-1 α transcription factor, complement triggering, integrin family, cell surface interactions, platelet signaling, VEGF signaling, thrombin /protease-activated receptor signaling pathway etc. Significant unique pathways with only hypo-methylated genes in SL-VTE included NOS activation and regulation, heme biosynthesis, NCAM1 inteactions, platelet degranulation and response to elevated platelet cytosolic Ca^2+^.On the other hand, uniquely hypermethylated pathways included Ca^2+^ activated K^+^ channels, extrinsic pathway, fatty acid activation, FGFR ligand binding and activation, formation of fibrin clot, GP1b-IX-V activation signaling, inflammasomes, platelet activation, NLRP3 inflammasome etc (Figure 5). (Supplementary table 3).

### Gene Enrichment analysis of hypo-methylated and hyper-methylatedgenes

The differentially methylated gene lists obtained for sea levelVTE patients and high altitude VTE patients were subjected to gene ontology analysis using ShinyGOv.0.61 (http://bioinformatics.sdstate.edu/go/). Most significantly enriched biological pathways included cell communication, signa ltransduction, cell growth/maintenance, transport, metabolism, Immune response, cell adhesion, apoptosis, lipid metabolism, regulation of cell cycle etc. Topbiological processes enriched in HA-VTE and SL-VTE group are depicted in Figure 6. Enriched cellular component included Plasma membrane, Cytoplasm, nucleus, exosomes, cell surface, centrosome, mitochondrian, actin cytoskeleton, lysosome, Voltage-gated potassium channel complex etc.; whereas molecular function included Cell adhesion molecule activity, Transporter activity, G-protein coupled receptor activity. Transcription factor activity, Receptor signaling complex scaffold activity, Voltage-gated ion channel activity, Oxido reductase activity, Calcium ion binding, Cyto skeletal anchoring activity Protein serine/threonine kinase activity.

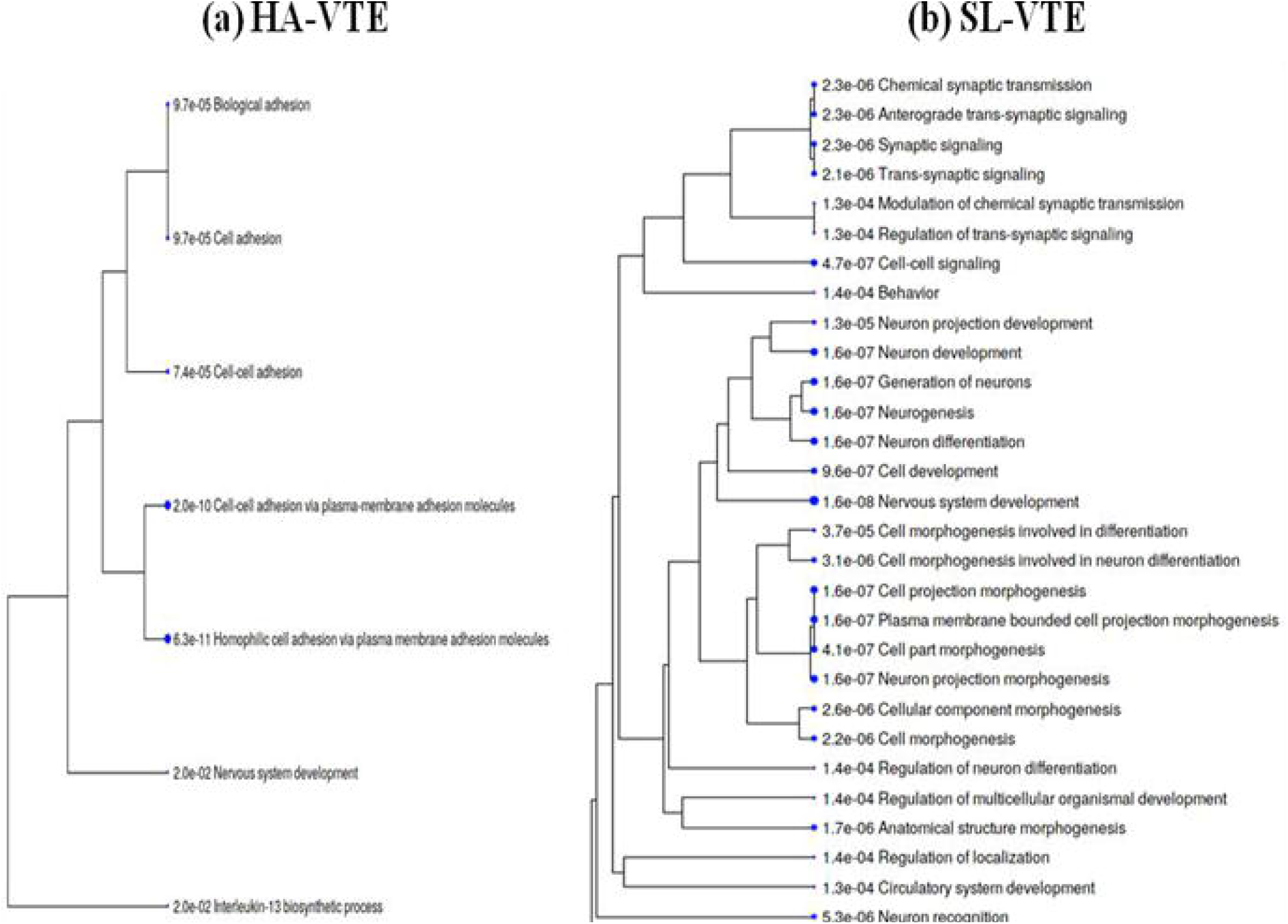

## Discussion

Aberrant DNA methylation on promoter affects gene expression and hinders promoter accessibility, and has been linked to multiple pathologies like variety of cancers, skin diseases, and cardiovascular diseases (Jelinek et al., 2011, Wang et al. 2018). Increased risk of thrombotic complications in high altitude climbers has been well documented. High altitude environment resulting in a hyper coagulabe state, is considered as an independent risk factor for VTE development (Kumar 2006, Damodar et al. 2018, Dutta et al. 2018). The rear every limited studies so far on DNA methylation in humans at high altitude (Childebayeva et al. 2019, Basak et al. 2017). Also, very scanty literature is available on methylation change during VTE event and blood coagulation (Aissi et al.2014, Ward-caviness et al. 2018, Noro et al. 2019). Global DNA methylation signatures in HA-VTE have not been studied so far! Infact, exact etiopathology of VTE inhumans in terms of epigenetic modulation (methylation) has not been elucidated. There are two separate linesof evidence; one is that hypoxic environment induces hyper-coagulation and another proves that hypoxia is associated with DNA methylation. However despite this, the interrelated studies on role of hypoxia induced DNA methylation changes in human VTE patients have not been documented so far. In the present study, we attempted to understand the role of methylation in high altitude hypoxia induced VTE.

The acute response to hypoxia involves changes in homeostatic regulation which is inturn controlled by large number of genes. Epigenetic modifications are plastic in nature and occur in response to the change in the environmental conditions (Bollati and Baccarelli, 2010). The present study utilizes gold standard method of sodium bisulphate sequencing technique to decipher epigenetic methylation changes during HA-VTE event.

### Differential methylation (DM) in High altitude VTE patients and Seal level VTE patients Hypo methylationin high altitude patients

Component of hypo methylation in high altitude patients compared to sea level patients included genes involved in cell adhesion and regulation, cell adhesion molecule 1 (CADM1), receptor-type tyrosine-protein phosphatase kappa (PTPRK); platelet derived growth factor (PDGFA),inflammation and immune response, C-X-C motif chemokine ligand 12 (CXCL12), immune surveillance, interleukin 18 (IL18), interleukin 4 (IL4), interferon regulatory factor 4 (IRF4) and neucleotide-binding oligomerization domain, leucine rich repeat and pyrin domain containing 1(NLRP1). Inflammation is reportedly higher upon exposure to high altitude (Hartman etal.2000) and platelets play a very important role in vascular homeostasis (Gupta et al. 2020), the hypo methylated (over-expressed) genes of inflammation and platelets activity may play an important role in predisposition towards HA-VTE. Apart from these, other hypo methylated genes in HAP were Proteasome 26S Subunit, Non-ATPase 14 (PSMD14) and Cullin 1 (CUL1) involved in ubiquitination and degradation of intra cellular proteins involved in cell cycle regulation and signal transduction and MutS Homolog 2 (MSH2) involved in DNA mismatch repair. Hypomethylated genes in HAP in comparison to HAC revealed oxygen-dependent coproporp hyrinogen-III oxidase, mitochondrial gene (CPOX), involved in biosynthetic pathway of heme, which might be in response to hypoxic conditions (Grek et al. 2011); N-Myc downstream-regulated gene 1 protein (NDRG1) which belongs to alpha/beta hydrolase and isreportedly involved in cell growth, immune and stress response and also negatively correlated with cancer progression (Fangetal. 2014, Kovacevic et al. 2016); and sphingomyelin synthase 1(SGMS1). The differential methylation in the said genes was mostly observed in the intergenic region and intronic regions, except IL18 with methylation in promoter region.

### Hyper methylation in High altitude patients

Hyper methylated genes in HAP in comparison to SLP included a transcription factor (GSC), Ribosomal protein S6 kinase alpha1 gene (RPS6KA1), which is a serine/threonine-protein kinase which signals and mediates mitogenic and stress induced activation of transcription factor. Amongst the other hyper-methylated genes were, transforming growth factor beta-2 (TGFB2), trans membrane receptor (NOTCH2), potassium voltage gated channel (KCND2), Toll-like receptor (TLR4) which activates intracellular signaling of NFκB, Rho-associated protein kinase (ROCK1) which is a key regulator of actin–myosin contraction, stability and cell polarity and zinc finger protein (ZFYVE16). Other hyper-methylated genes in high altitude patients in comparison to respective controls (HAC) were cell adhesion molecule (Cadherin, CDH5), ETS domain transcription factor (ELF1), Kalirin (KALRN), which is involved in inducing various signaling mechanisms, Neuro trophic tyrosine kinase, receptor type2 (NTRK2), Srckinase (SKAP2), which plays a role in activation of immune system, t-SNARE (SNAP25) involved in molecular regulation of neuro transmitter release and trans membrane serine protease (TMPRSS2) involved in tissue remodeling, blood coagulation and inflammatory response etc. Except for the KALRN, all other hyper methylated genes in this category showed DM in intergenic and intronic regions.

### Hypo methylation in sea level patients

The percentage of DM was much higher in case of sea level patients compared to high altitude patients. The differentially methylated genes belonged to various biological pathways (as mentioned in supplementary table 3), we have enlisted 41 hypo-methylated and 76 hyper-methylated genes amongst them which were found to be significantly involved in multiple pathways.

Amongst the 41 hypo methylated genes, a large number of genes belonged to (1) immune response pathway such as ELMO1 (engulfment and cell motility protein 1), IL18 (interleukin 18,apro-inflmmatory cytokine involved in T-helper cell, natural killer cell and cell immune response), MBL2 (mannose Binding lectin 2, an important element of innate immune response), RIPK2 (receptor-interacting serine/threonine-protein kinase involved in innate and adaptive immune response), ICOS (Inducible T-cell co-stimulator, an immune check point protein belonging to CD28 and CTLA-4 cell surface receptor family); (2) Transcriptional activation like BCL9 (B-cell lumphoma 9, a transcriptional co-activator), CREM (cAMP responsive element modulator which is involved in transcriptional response to stress, MDFIC (MyoD family inhibitor domain-containing protein, a transcriptional activator of repressor), MYOCD (Myo Cardin), KDM3A (Lysine demethylase 3A, has DNA binding transcription factor activity), TCF7L2 (transcription factor 7 like 2, plays a key role in Wnt signaling pathway and implicated in blood glucose homeostasis, RUNX1 (Runt-related transcription factor for development of normal hematopoiesis; (3) genes involved in cell/ platelet adhesion such as CD226 (cell adhesion molecule mediating adhesion of platelets and mega karyocytes to vascular endothelial cells), DCC (Deleted in colorectal carcinoma, a trans membrane protein and member of immunoglobulin super family of cell adhesion molecules, ST8S1A4 (Sialyl transferase, modulator of adhesive properties on nCAM1); (4) membrane receptors and regulatory proteins viz., CXCR4(C-X-C motif chemokine receptor 4, a G-protein coupled receptor for protein ubiquitination), DUSP6 (Dual specificity phosphatase 6), GNAL (G-protein alpha subunit), GRIP2 (Glutamate receptor interaction protein 2), PPP3R1 (protein phosphatase 3 regulatory subunit B, alpha, calcium stimulated) and PRKAR1B (cAMP dependent protein kinase type 1-beta regulatory protein, involved in cAMP signaling in cells). Besides these major sub classes, other differentially hypo methylated genes in this category included those involved in maintaining cyto skeletal integrity such as COL11 A1 (Collagen type XI A1 chain, a constituent of extracellular matrix structure and playing an important role in fibrillogenesis by controlling growth of collagen fibrils) and TLN1(Talin1, forming connection of major cyto skeletal structures to the plasma membrane); channel proteins such as KCNC2 (voltage gated potassium channel) and TRPC7 (Transient receptor potential cation channel subfamily, member 7) and genes involved in oxidative phosphorylation like NDUFAF2 (NADPH: ubiquinone oxireductase complex assembly factor 2, which acts as molecular chaperon for mitochondria complex I and has role in electron transfer from NADH to the respiratory chain and CPOX (Copropor phyrinogen-III oxidase, which is a mitochrondrial enzyme involved in production of heme molecule). Histidine-rich glycoprotein (HRG) gene was also found to be hypo methylated. This genes has diverse functions, it binds to number of ligands such as heme, heparin, thrombospondin, plasminogen etc and regulates processes such as immune system, pathogen clearance, cell adhesion, angiogenesis, coagulation and fibrinolysis. Another interesting hypo-methylated gene was IQGAP1 (IQ Motif containing GTPase activating protein1), which interacts with components of cytoskeletal system, cell adhesion molecules and also with several signaling molecules to regulate cell morphology and motility. KDM3 A gene was hypo methylated in exonic region, CD226 in promoter region and all others in respective intergenic and intronic regions.

### Hype rmethylation in sea level patients

Several transcription factors were hyper-methylated in sea level patients such as BCL3 (B-cell lymphoma 3 encoded protein, a transcriptional co-activator), DDX58 (Dead box family of RNA helicases, a transcriptional repressor), ETV1 (ETS variant transcription factor 1), PBX1 (pre-B-cell leukemia transcription factor 1), TLE1 (transducin-like enhancer protein 1, a transcriptional co-repressor that binds to a number of transcription factors like NFκB) and SPI1 (transcription factor PU.1, a transcriptional activator involved in activation of macrophagesor B-cells). Although increased platelet activity and fibrinogen levels collectively account for pro-thrombotic state (Kotwaletal.2007), many genes involved in platelet activation and coagulation pathway showed hyper methylation (down-regulation) including CD36 (collagen type1 receptor or thrombospondin receptor, a major glycoprotein of platelet surface), F3, also known as tissue factor (TF) which facilitates blood coagulation by forming a complex with circulating factor VIIor VIIa, which in turn activates factor X (Owens et al. 2010), PDGFA and PDGFRA (Platelet-derived growth factor -subunit A and –receptor alpha, respectively), TFPI (tissue factor pathway inhibitor, a kunitz-type serine protease inhibitor that regulates TF-dependent blood coagulation) and MASP1 (Mannose-binding lectin associated serine protease 1) that functions as a component of lectin pathway of complement activation, complement pathway and plays an essential role in immune response and coagulation) Hyper methylation in tissue factor F3 is somehow counter balanced by hyper methylation in TFPI. Hypermethylated genes SLP also included genes encoding for proteins involved in cell adhesion or focal adhesion such as ARAP2 (ArfGAP with Rho GAP domain), CD44 (a cell surface glycoprotein), CDH2 (Cadherin 2, another cell surface glycoprotein), DSG1 (Desmoglin 1-cadherin-like transmembrane glycoprotein that form a major component of desmosomes), LPP (Lipoma-preferred partner, playing a structural role at the sites of cell adhesion thus maintain cell shape and moility), PAK1 (Serine/threonine kinase, involved in intracellular signaling and plays an important role in cytoskeleton dynamics and cell adhesion)and SORBS1 (Sorbin 1 and SH3 domain containing protein involved in formation of actin stress fibers and focal adhesion). Hyper methylated growth factors included FGF10 (fibroblast growth factor10), GAB2 (growth factor receptor bound protein 2), GF1(growth factor independence1, a transcriptional repressor), IGF1(insulin like growth factor 1), IGF1R (insulin like growth factor receptor),TGB1 (transforming growth factor beta1),TGFBR3 (transforming growth factor beta receptor 3). Hyper methylation of genes involved in immune-regulatory and inflammatory processes included CCL26 (C-C motif chemokine lignad L6), CD200R1 (C-C motif chemokine liand L6) and PRDM1 (PR domain zinc finger protein 1, a transcription factor that mediates innate and adaptive immune response). Other genes of interest in this category included EDN1 (endothelin 1), which is an endothelium derived vasoconstrictor and EGLN3(Prolyl hydroxylase), which is an important isozyme in limiting physiological activation of Hypoxia inducible factor (HIF) in hypoxia. Besides DDX58 gene which showed hypermethylationin 3’-region, all others were hypermethylated in intronic and intergenic regions.

Our findings give global DNA methylation patterns in Indian population, in an event of venous thrombosis for the first time, both at high altitude as well as sea level. Further studies with more number of samples would helpful in validating and generalizing present findings.

## Data Availability

All data produced in the present work are contained in the manuscript

## Summary

To the best of our knowledge, hypoxia as encountered at high altitude and associated venous thrombosis phenotype, has never been discussed and appreciated in perspective of DNA methylation. Present study identified differential hypo methylation in cell adhesion and inflammation genes and hypermethylation of certain transcription factors and channel proteins in high altitude VTE patients. A distinct pattern of methylation was observed in HA-VTE patients and SL-VTE patients. Overall, the work primarily provides robust at a demonstrating that hypoxia reinforces global methylation events.

## Limitation of the study

Blood samples from subjects with mixed ethnic backgrounds, have been collected at different time points, as and when available. The methylation pattern of an individual changes with change in altitude, environment and even lifestyle habits. In the present study, these factors have not been taken into account. Also, due to technical reasons/sample unavailability, we could study only a limited number of subjects in each group.

## Acknowledgement

The authors are extremely thankful to all the Indian Army volunteers who participated in the study. We are also grateful to our co-investigators at Army R & R hospital, Delhi and Western command hospital, Chandimandir, who facilitated us with sample collection. We express our sincere thanks to ex Director, DIPAS, Dr. Rajiv Varshney, for logistic support and guidance.

## Author’sContribution

SS: Performed the Experiments, analyzed data and wrote the manuscript, IG: Manuscript writing and Editing BK: Assisted in performing the experiments and sample collection, UY and JS: Collection of samples, L Gand RV: Logistic support and manuscript proof reading.

