## Supplementary Tables for "Identification of Global DNA Methylation Signatures in Patients of High Altitude Induced Venous Thrombo-Embolism (HA-VTE)"

***Table1:*** Differentially methylated pathways in High Altitude Patients in comparison to Sea level patients

| **CommonPathways** | **Hypo-methylated** | **Hyper-methylated** | | |
| --- | --- | --- | --- | --- |
| AP-1transcriptionfactornetwork | CXCL12;IRF4;PTPRK;CUL1; | GSC;ZFYVE16;TGFB2; | | |
| Apoptosis | PSMD14;PPP3R1; | ROCK1; | | |
| Beta1 integrin cell surface  interactions | IRS2;CXCL12;IL4R;IRF4;MSH2;  PTPRK;ITSN1;CUL1; | GSC; RPS6KA1;  ROCK1; | ZFYVE16; | TGFB2; |
| Endothelinsignalingpathway | IRS2;CXCL12;IL4R;IRF4;MSH2;  PTPRK;ITSN1;CUL1; | GSC; RPS6KA1;  ROCK1; | ZFYVE16; | TGFB2; |
| IGF1pathway | IRS2;CXCL12;IL4R;IRF4;MSH2;  PTPRK;ITSN1;CUL1; | GSC; RPS6KA1;  ROCK1; | ZFYVE16; | TGFB2; |
| ImmuneSystem | IRS2;IL18;PSMD14;IRF4;NLRP1;  CUL1; | RPS6KA1;TLR4; | | |
| Notch signalingpathway | CUL1; | NOTCH2; | | |
| PDGFreceptorsignalingnetwork | IRS2;CXCL12;IL4R;IRF4;PDGFA;  MSH2;PTPRK;ITSN1;CUL1; | GSC; RPS6KA1;  ROCK1; | ZFYVE16; | TGFB2; |
| Signaling events mediated by  VEGFR1andVEGFR2 | IRS2;CXCL12;IL4R;IRF4;MSH2;  PTPRK;ITSN1;CUL1; | GSC; RPS6KA1;  ROCK1; | ZFYVE16; | TGFB2; |
| Thrombin/protease-activated  receptor(PAR)pathway | IRS2;CXCL12;IL4R;IRF4;MSH2;  PTPRK;ITSN1;CUL1; | GSC; RPS6KA1;  ROCK1; | ZFYVE16; | TGFB2; |
| Urokinase-type plasminogen  activator (uPA) and uPAR-mediated signaling | IRS2;CXCL12;IL4R;IRF4;MSH2;PTPRK;ITSN1; CUL1; | GSC; RPS6KA1;ROCK1; | ZFYVE16; | TGFB2; |
| **Significantuniquepathways** | **Hypo-methylated** |  | | |
| Adherensjunctionsinteractions | CADM1; | _ | | |
| Biologicaloxidations | CYP4F11; | _ | | |
| EPOsignalingpathway | IRS2; | _ | | |
| HIF-1-alpha transcription factor  network | CXCL12; | _ | | |
| Inflammasomes | NLRP1; | _ | | |
| p53pathway | MSH2; | _ | | |
| p53-Independent DNA Damage  Response | PSMD14; | _ | | |
| **Significantuniquepathways** |  | **Hyper-methylated** | | |
| PotassiumChannels | _ | KCND2; | | |
| TollReceptorCascades | _ | RPS6KA1;TLR4; | | |

***Table2 :*** Differentially methylated pathways in High Altitude Patients in comparison to High altitude controls

| **Common Pathways** | **Hypo-methylated** | **Hyper-methylated** |
| --- | --- | --- |
| AP-1transcriptionfactornetwork | NDRG1; | ELF1;TMPRSS2; |
| Beta1integrincellsurfaceinteractions | NDRG1; | ELF1; KALRN; TMPRSS2;NTRK2; |
| EGFR-dependent Endothelin signaling events | NDRG1; | ELF1;KALRN;TMPRSS2;  NTRK2; |
| Endothelin signaling pathway | NDRG1; | ELF1;KALRN;TMPRSS2;  NTRK2; |
| Integrin family cell surface interactions | NDRG1; | ELF1;KALRN;TMPRSS2;  NTRK2; |
| PDGFR-beta signaling pathway | NDRG1; | ELF1;KALRN;TMPRSS2;  NTRK2; |
| Signaling events mediated by focal adhesion kinase | NDRG1; | ELF1;KALRN;TMPRSS2;  NTRK2; |
| Urokinase-type plasminogen activator(uPA) and uPAR-  Mediated signaling | NDRG1; | ELF1;KALRN;TMPRSS2;  NTRK2; |
| VEGF and VEGFR signaling network | NDRG1; | ELF1;CDH5;KALRN;  TMPRSS2;NTRK2; |
| Thrombin/protease-activated receptor(PAR)pathway | NDRG1; | ELF1;KALRN;TMPRSS2;  NTRK2; |
| **Significant unique pathways** | **Hypo-methylated** |  |
| Heme biosynthesis | CPOX; |  |
| HIF-1-alphatranscriptionfactornetwork | NDRG1; |  |
| Metabolism and lipoproteins of lipids | SGMS1; |  |
| **Significant unique pathways** |  | **Hyper-methylated** |
| Angiopoietin receptor Tie2-mediated signaling |  | ELF1; |
| E-cadherin signaling events |  | TMPRSS2;NTRK2; |
| GABA synthesis release, re-uptake and degradation |  | SNAP25; |
| Cell-Cell communication |  | CDH5;SKAP2; |

***Table3:*** Differentially methylated pathways in Sea level Patients in comparison to Sea level controls

| **Common Pathways** | | **Hypo-methylated** | | **Hyper-methylated** |
| --- | --- | --- | --- | --- |
| Adaptive Immune System | | ICOS;RIPK2;CD226; | | ITGB1;PSMB1;CD200R1;PAK1;MAP3K7;  PTPRC; |
| AP-1transcriptionfactornetwork | | CREM; MDFIC; NEDD4L;PPARGC1A; TCF7L2; RUNX1;ZFYVE16;CXCR4;KDM3A;  BCL9; | | IL10; MAP2K4; TLE1; ETV1; EGLN3; PAK1;DUSP16; GAB2; TGFBR3; MAP3K7; YWHAH;PBX1;EDN1;DCN;SRC;PDGFRA;SPI1;GZMB; |
| Apoptosis | | DCC;PPP3R1; | | PSMB1;FADD;PKP1;DSG1;DAPK1;GZMB; |
| Beta1integrincellsurfaceinteractions | | IQGAP1; NDUFAF2; CREM;MYOCD; MDFIC; NEDD4L;MSH2;PPARGC1A;TCF7L2;DUSP6; ELMO1; RIPK2;SPRY2; COL11A1; RUNX1;ZFYVE16; CXCR4; KDM3A;PRKAR1B;TLN1;BCL9; | | ITGB1; ROBO1; ERC1; RBL2; IL10; IGF1R;MAP2K4; LAMA2; TLE1; IGF1; ETV1; KALRN;DIAPH3;GNA15;EGLN3;PRDM1;PAK1;DUSP16;GAB2; LPP; INSR; TGFBR3; MAP3K7; YWHAH;PBX1; EDN1; DCN; SORBS1; SRC; CCL26;STXBP4; PDGFRA; SPI1; PTPRC; BCL3; OPRM1;ARAP2;CDH2; GZMB; |
| Complement cascade | | MBL2; | | MASP1; |
| Endothelin signaling pathway | | IQGAP1; NDUFAF2; CREM;MYOCD; MDFIC; NEDD4L;MSH2; PPARGC1A; ADCY8;TCF7L2; DUSP6; ELMO1;RIPK2;SPRY2;RUNX1;GNAL;ZFYVE16; CXCR4; KDM3A;TLN1;BCL9; | | ITGB1; ROBO1; ERC1; RBL2; IL10; IGF1R;MAP2K4; TLE1; IGF1; ETV1; KALRN; DIAPH3;GNA15; EGLN3; PRDM1; PAK1; DUSP16; GAB2;LPP; INSR; TGFBR3; MAP3K7; YWHAH; PBX1;EDN1; PLCB1; DCN; SORBS1; SRC; CCL26;STXBP4; PDGFRA; SPI1; PTPRC; BCL3; OPRM1;ARAP2;CDH2; GZMB; |
| FGF signaling pathway | | SPRY2; | | SRC;CDH2; |
| GABA receptor activation | | ADCY8;GNAL; GABRB2; | | GABRG2;GNGT1;GABBR2;GABBR1;GNG4;GABRG3; |
| Hemostasis | | HRG; KIFAP3; WEE1; TRPC7;PRKAR1B; GRIP2; TLN1;ABCC4; | | ITGB1; CD36; LRRC16A; GNA15; GUCY1A2; F3;DGKD;SRC; TFPI; |
| Heparan sulfate biosynthesis | | HS3ST3A1; | | NDST3; |
| HIF-1-alphatranscriptionfactor  Network | | CXCR4; | | EGLN3;EDN1; |
| Immune System | | ICOS; RIPK2; SPRY2; CD226;IL18;SP100;TLN1;MBL2; | | ITGB1;PSMB1;CD200R1;MAP2K4;FADD;PAK1;GAB2; MAP3K7; SRC; MASP1; PANX1; CD44;PTPRC;DDX58; |
| Initial triggering of complement | | MBL2; | | MASP1; |
| Innate Immune System | | RIPK2;MBL2; | | MAP2K4;FADD;MAP3K7;MASP1;PANX1;  DDX58; |
| Integrin family cell surface interactions | | IQGAP1; NDUFAF2; CREM;MYOCD; MDFIC; NEDD4L;MSH2;PPARGC1A;TCF7L2;DUSP6; ELMO1; RIPK2;SPRY2; COL11A1; RUNX1;ZFYVE16; CXCR4; KDM3A;PRKAR1B;TLN1;BCL9; | | ITGB1; ROBO1; ERC1; RBL2; IL10; IGF1R;MAP2K4; LAMA2; TLE1; IGF1; ETV1; KALRN;DIAPH3;GNA15;EGLN3;PRDM1;PAK1;DUSP16;GAB2; LPP; INSR; TGFBR3; MAP3K7; YWHAH;PBX1; EDN1; DCN; SORBS1; SRC; CCL26;STXBP4; PDGFRA; SPI1; PTPRC; BCL3; OPRM1;ARAP2;CDH2; GZMB; |
| Nectin adhesion pathway | | IQGAP1; NDUFAF2; CREM;MYOCD; MDFIC; NEDD4L;MSH2; PPARGC1A; TCF7L2;DUSP6; ELMO1; RIPK2;SPRY2; RUNX1; ZFYVE16;CXCR4;KDM3A;TLN1;BCL9; | | ITGB1; ROBO1; ERC1; RBL2; IL10; IGF1R;MAP2K4; TLE1; IGF1; ETV1; KALRN; DIAPH3;GNA15; EGLN3; PRDM1; PAK1; DUSP16; GAB2;LPP; INSR; TGFBR3; MAP3K7; YWHAH; PBX1;EDN1; DCN; SORBS1; SRC; CCL26; STXBP4;PDGFRA; SPI1; PTPRC; BCL3; OPRM1; ARAP2;CDH2;GZMB; |
| PDGF receptor signaling network | | IQGAP1; NDUFAF2; CREM;MYOCD; MDFIC; NEDD4L;MSH2; PPARGC1A; TCF7L2;DUSP6; ELMO1; RIPK2;SPRY2; RUNX1; ZFYVE16;CXCR4;KDM3A;TLN1;BCL9; | | ITGB1; ROBO1; ERC1; RBL2; IL10; IGF1R;MAP2K4; TLE1; IGF1; ETV1; KALRN; DIAPH3;GNA15; EGLN3; PRDM1; PAK1; DUSP16; GAB2;LPP;INSR;PDGFA; TGFBR3;MAP3K7;YWHAH;PBX1; EDN1; DCN; SORBS1; SRC; CCL26;STXBP4;PDGFRA;SPI1;PTPRC;BCL3;OPRM1;  ARAP2;CDH2;GZMB; |
| Platelet activation, signaling and aggregation | | TRPC7;GRIP2;TLN1;ABCC4; | | GNA15;DGKD;SRC; |
| Platelet Aggregation (Plug Formation) | | TLN1; | | SRC; |
| Platelet homeostasis | | TRPC7; | | GUCY1A2; |
| Signaling by EGFR | | ADCY8;SPRY2;PRKAR1B; | | SRC; |
| Signaling by FGFR | | ADCY8;SPRY2;PRKAR1B; | | SRC;FGF10; |
| Signaling events mediated by focal adhesion kinase | | IQGAP1; NDUFAF2; CREM;MYOCD; MDFIC; NEDD4L;MSH2; PPARGC1A; TCF7L2;DUSP6; ELMO1; RIPK2;SPRY2; RUNX1; ZFYVE16;CXCR4;KDM3A;TLN1;BCL9; | | ITGB1; ROBO1; ERC1; RBL2; IL10; IGF1R;MAP2K4; TLE1; IGF1; ETV1; KALRN; DIAPH3;GNA15; EGLN3; PRDM1; PAK1; DUSP16; GAB2;LPP; INSR; TGFBR3; MAP3K7; YWHAH; PBX1;EDN1; DCN; SORBS1; SRC; CCL26; STXBP4;PDGFRA; SPI1; PTPRC; BCL3; OPRM1; ARAP2;CDH2;GZMB; |
| Signaling events mediated by VEGFR1 and VEGFR2 | | IQGAP1; NDUFAF2; CREM;MYOCD; MDFIC; NEDD4L;MSH2; PPARGC1A; TCF7L2;DUSP6; ELMO1; RIPK2;SPRY2; RUNX1; ZFYVE16;CXCR4;KDM3A;TLN1;BCL9; | | ITGB1; ROBO1; ERC1; RBL2; IL10; IGF1R;MAP2K4; TLE1; IGF1; ETV1; KALRN; DIAPH3;GNA15; EGLN3; PRDM1; PAK1; DUSP16; GAB2;LPP; INSR; TGFBR3; MAP3K7; YWHAH; PBX1;EDN1; DCN; SORBS1; SRC; CCL26; STXBP4;PDGFRA;SPI1;PTPRC;BCL3;OPRM1;ARAP2;  CDH2;GZMB; |
| Thrombin/protease-activated receptor(PAR)pathway | | IQGAP1; NDUFAF2; CREM;MYOCD; MDFIC; NEDD4L;MSH2; PPARGC1A; TCF7L2;DUSP6; ELMO1; RIPK2;SPRY2; RUNX1; ZFYVE16;CXCR4;KDM3A;TLN1;BCL9; | | ITGB1; ROBO1; ERC1; RBL2; IL10; IGF1R;MAP2K4; TLE1; IGF1; ETV1; KALRN; DIAPH3;GNA15; EGLN3; PRDM1; PAK1; DUSP16; GAB2;LPP; INSR; TGFBR3; MAP3K7; YWHAH; PBX1;EDN1;PLCB1; DCN; SORBS1;SRC; CCL26;  STXBP4; PDGFRA; SPI1; PTPRC; BCL3; OPRM1;ARAP2;CDH2; GZMB; |
| Urokinase-type plasminogen activator (uPA) and uPAR-mediated signaling | | IQGAP1; NDUFAF2; CREM;MYOCD; MDFIC; NEDD4L;MSH2; PPARGC1A; TCF7L2;DUSP6; ELMO1; RIPK2;SPRY2; RUNX1; ZFYVE16;CXCR4;KDM3A;TLN1;BCL9; | | ITGB1; ROBO1; ERC1; RBL2; IL10; IGF1R;MAP2K4; TLE1; IGF1; ETV1; KALRN; DIAPH3;GNA15; EGLN3; PRDM1; PAK1; DUSP16; GAB2;LPP; INSR; TGFBR3; MAP3K7; YWHAH; PBX1;EDN1; DCN; SORBS1; SRC; CCL26; STXBP4;PDGFRA; SPI1; PTPRC; BCL3; OPRM1; ARAP2;CDH2;GZMB; |
| Voltage gated Potassium channels | | KCNC2;KCND2; | | KCNB2;KCNA6;KCNA1;KCNV1;KCNQ1; |
| **Significant unique pathways** | | **Hypo-methylated** | |  |
| eNOS activation and regulation | | GCH1; | | _ |
| Heme biosynthesis | | CPOX; | | _ |
| NCAM1interactions | | ST8SIA4; | | _ |
| Platelet de-granulation | | TLN1;ABCC4; | | _ |
| Regulation of Water Balance by Renal Aquaporins | ADCY8;PRKAR1B; | | _ | |
| Response to elevated platelet cytosolic Ca2+ | GRIP2;TLN1;ABCC4; | | _ | |
| **Significant unique pathways** |  | | **Hyper-methylated** | |
| Ca2+activated K+ channels | _ | | KCNN3; | |
| Extrinsic Pathway | _ | | F3;TFPI; | |
| Extrinsic Pathway for Apoptosis | _ | | FADD; | |
| Fatty acid activation | _ | | ACSL1; | |
| FGFR ligand binding and activation | _ | | FGF10; | |
| Formation of Fibrin Clot (Clotting Cascade) | _ | | F3;TFPI; | |
| G beta: Gamma signaling through PLC beta | _ | | PLCB1; | |
| GP1b-IX-Vactivationsignaling | _ | | SRC; | |
| Inflammasomes | _ | | PANX1; | |
| Platelet Adhesion to exposed  Collagen | _ | | ITGB1;CD36; | |
| Respiratory electron transport, ATP synthesis by chemiosmotic coupling, and heat production by uncoupling proteins. | _ | | NDUFA4;NDUFS4; | |
| The citric acid(TCA) cycle and respiratory electron transport | _ | | NDUFA4;NDUFS4; | |
| NLRP3 inflammasome | _ | | PANX1; | |
| Thrombin signalling through proteinase activated receptors  (PARs) | _ | | GNA15;SRC; | |
| VEGF ligand-receptor interactions | _ | | NRP1; | |
